# Outcomes of gastrointestinal fistulas: Results from a multi-continent, multi-national, multi-center cohort

**DOI:** 10.1101/2021.09.23.21262745

**Authors:** Humberto Arenas Márquez, María Isabel Turcios Correia, Juan Francisco García, Roberto Anaya Prado, Arturo Vergara, Jorge Luis Garnica, Alejandra Cacho, Daniel Guerra, Miguel Mendoza Navarrete, Sergio Santana Porbén

**Affiliations:** Unidad de Falla intestinal SANVITE, Hospital San Javier, Avenida Pablo Casals 640, Prados Providencia, Guadalajara 44670, Estado de Jalisco, Estados Unidos Mexicanos

**Author notes:** Address all correspondence to: Humberto Arenas Márquez, Electronic address.

**Keywords:** Gastrointestinal fistula, Cohort, Mortality, Survival, Hospitalization, Spontaneous closure

## Abstract

**Rationale:** Gastrointestinal fistulas (GIF) represent a severe and potentially lethal complication of the hospital surgical activity. However, evidences are lacking about prognosis and outcomes of GIF in Latin America (LATAM) hospitals.

**Objective:** To describe the prognosis and outcomes GIF in LATAM hospitals.

**Study design:** Prospective, longitudinal, cohort-type study. The cohort fostered three cross-sectional examinations: First examination: On admission of the patient in the study; Second examination: Thirty days later; and Third (and last) examination: Sixty days after patient’s admission.

**Study serie:** One hundred seventy-seven patients (*Males*: 58.2 %; *Average age*: 51.0 ± 16.7 years; Ages ≥ 60 years: 36.2 %) diagnosed with, and assisted for, GIF (ECF: Enterocutaneous: 64.9 % *vs*. EAF: Enteroathmospheric: 35.1 %) in 76 LATAM hospitals (13 countries) and Europe (4).

**Methods:** Condition (Alive *vs*. Deceased) and hospital status (Hospitalized *vs*. Discharged) of the patient, and the GFI patency (Closed *vs*. Non closed) were recorded in each of the cohort’s examination. Indicators of GFI prognosis thus constructed were correlated demographical, sanitary, surgical and nutritional characteristics of the patients

**Results:** On conclusion of the study indicators of GIF prognosis behaved as follows: *Mortality*:14.7 %; *Prolonged hospitalization*: 47.4 %; *Spontaneous closure of GIF*: 36.2 %. Type of GIF influenced upon patient’s survival: *ECF*: 87.0 % vs. *EAF*: 82.3 % (Δ = +4.7 %; *χ*2 = 6.787; p < 0.05). In each examination of the cohort, the number of surviving subjects was always greater among those with ECF: *After 30 days*: ECF: 92.1 % vs. EAF: 83.9 % (Δ = +8.2 %); *After 60 days*: ECF: 98.1 % vs. EAF: 90.4 % (Δ = +7.7 %; *χ*2 = 13.764; p < 0.05). On the other hand, hospital stay was prolonged in the subjects of elective surgery (*Elective surgery*: 61.4 % vs. *Emergency surgery*: 38.3 % (Δ = +23.1 %; *χ*^2^ = 9.064; p < 0.05) and those with a reduced calf circumference (*χ*^2^ = 12.655; p < 0.05). Location of the fistula also influenced upon prolongation of hospital stay (*χ*^2^ = 7.817; p < 0.05).

**Conclusions:** Type of GIF influences upon survival of the patient. On the hand, hospital stay was dependent upon type of surgery previously performed, location of the fistula, and calf circumference value on admission in the study serie.

## INTRODUCTION

Gastrointestinal fistulas (GIF) represent a serious complication of the surgical processes, and bear a high risk of hydroelectrolyte disorders, sepsis, malnutrition and death.^1-2^ Hence, with repercussions of GIF known, timely recognition, accurate diagnosis (integrating criteria on the spontaneous closure-also read as conservative | non-surgical-of the GIF); and adoption of the required measures for hydroelectrolyte repletion, nutritional support and containment of sepsis are imperative for a better prognosis of GIF.

Incidence of GIF is estimated to vary between 4 – 20 % of the operated patients,^1-2^ but this estimate might vary with the typology of the health institution, and the volume and complexity of the surgeries completed in a year. Almost half of the GIF originate in patients in whom no intestinal anastomosis is performed, but suffer inadvertent enterectomies during the surgical act.^3-4^ The other half of GIF is caused by disruption (partial or complete) of the intestinal suture made. Reoperations with extensive lysis of adhesions and adherences, laparoscopic surgery, reparation of ventral hernias, tumor cytoreduction, and trauma inflicted during surgery not related with tumor cytoreduction are among the surgical procedures commonly associated with postsurgical GIF.^3-4^ GIF-related mortality could be as high as 80 % of the affected patients.^3-4^ It is then immediate that management of GIF leads to prolonged hospitalizations, a higher quota of clinical and surgical actions, and increased health care costs.

Management and resolution of GIF have called the attention of groups of experts and professional societies alike. In this regard, the “ASPEN/FELANPE clinical guidelines for nutrition support of adult patients with enterocutaneous fistula” are to be mentioned.^5-6^ Identification of the origin of the fistula, containment of surgical damage, and exteriorization of the fistula leakage might be actions initially recommended. Further actions in the management of GIF might imply the better assessment on their likely spontaneous closure. It is worthy to note coming to this point in the present narrative Crohn disease, malignancies of the gastrointestinal tract (GIT), and a hostile abdomen due to adherences, adhesions and reinterventions are always mentioned as negative predictors of the spontaneous closure of the fistula.

There are previous studies on the prevalence of GIF in hospitals of Latin America (LATAM), and practices adopted by the surgical care teams for the management and resolution of GIF. For the same reason, there are no estimates of the current effectiveness of different management guidelines that could be followed for GIF resolution.

The goals previously exposed might be achieved through “research outcomes”-type studies reuniting data generated by the surgical care teams managing GIF locally in order to show the current state of therapeutic effectiveness, to identify those actions leading to an unsatisfactory evolution of both the fistula and the patient, and, for the same reason, to find those teams distinguishing themselves for a higher therapeutic effectiveness for exploring the conducted practices, and validate and disseminate them to the others. Regarding this issue, the “Consenso Mexicano para el Tratamiento Integral de las Fistulas Digestivas” (first published 21 years ago) suggested the development, implementation and management of a national registry reuniting those variables that could be associated with adverse events during GIF management in order to correct inadequate practices and at the same improve quality of treatment.^7^ Recent advances in information and communication technologies have provided researchers with sophisticated tools for building and managing large databases and analyzing massive amounts of data, all of them eventually leading to a better treatment of GIF.

Given the aforementioned, the Federación Latinoamericana de Terapia Nutricional, Nutrición Clínica y Metabolismo^*^ has launched the “Fistula Day” Project as a multi-center, multi-national effort having as supraobjective the drafting and validation of the “Good Practices” in the management of GIF. Such supraobjective will be met by means of regular surveys among LATAM institutions dedicated to GIF management, and collection of relevant data on the clinical characteristics of the GIF patients, locally conducted practices, and results observed at the moment of the survey. With this purpose, FELANPE has commissioned the SANVITE Unit for Intestine Failure at “San Javier” Hospital (Guadalajara, Estado de Jalisco, México) for the design, conduction and management of activities revealing the current surgical treatment of GIF, the effectiveness of such treatments, and the nutritional care provided to the GIF patient as part of such treatments.

## PRESENTATION OF THE FISTULA DAY PROJECT

The “Fistula Day” is a research outcome-type project oriented to the construction of a map with the diagnosis, treatment and resolution of GIF in LATAM hospitals. As such, the “Fistula Day” foresees the completion of cross-sectional surveys at regular intervals among LATAM hospitals in order to obtain estimates of the operational characteristics of the hospitals treating GIF patients on one hand; and the demographic, sanitary, clinical-surgical and nutritional characteristics of the patients on the other; as well as the surgical practices conducted in them for intervening the surgical damage, and the nutritional support schemes currently administered to the patient as part of the resolution of GIF.

Integrating the results of the cross-sectional surveys completed as part of the “Fistula Day” into a cohort will also serve to assess the impact of findings and practices revealed upon 3 outcomes of interest for researchers: survival of the patient, prolongation of the hospital stay, and the spontaneous closure of the fistula. Eventually, the “Fistula Day” will provide the methodological and operational foundation for assessing the impact of the “Good Practices” ultimately adopted in the diagnosis, treatment and follow-up of GIF.

The design of the “Fistula Day” will be presented in this report, along with the demographical, sanitary, surgical and nutritional characteristics of the patients admitted in the first cohort, and the associations found with the indicators of prognosis and outcomes of GIF at the end of the research. Influence of the characteristics of the participating hospitals upon prognosis and outcomes of GIF will be addressed in an accompanying paper.

## MATERIAL AND METHOD

### Study design

Prospective, multi-national, multi-center cohort-type study. The design of the study foresaw 3 cross-sectional examinations in different times: 0, 30 and 60 days. Admission of the hospitals participating in the “Fistula Day” was completed on the first examination (Day 0). Successive examinations at 30 and 60 days were made for documenting the evolution of the patient, surgical practices performed, therapeutic response, and nutritional care administered.

### Inclusion criteria

Adults patients diagnosed and treated for GIF in second- and third-level LATAM hospitals. GIF was established after output of intestinal liquor through an orifice in the abdominal wall and/or the exposure of the intestinal mucosae to the exterior.^1-6^

### Exclusion criteria

Patients with fistulas of the biliary tree and the pancreatic duct were excluded.

### Methods

LATAM medical centers dedicated to the treatment of GIF were invited to participate in the “Fistula Day” through communications made by the societies, associations and colleges represented in FELANPE, notifications issued through social networks, and direct electronic messages. A web page^†^ was open for registering the participating center, downloading and management of “Fistula Day” tools and resources, and communication and interaction between researchers and participating centers.

As previously mentioned, the design of “Fistula Day” comprised 3 cross-sectional examinations. The first examination was completed on May 9th, 2018 with the registry of participating hospitals, and the collection of their sanitary and managerial characteristics. Once admission of the hospital in the “Fistula Day” was completed, patients locally assisted for GIF, and their demographical, clinical, surgical and nutritional characteristics were then entered into the forms provided by the design of the research. Current practices related with the surgical treatment of GIF, along with implemented nutritional support schemes, were also recorded.

Forms with data collected during the first examination were submitted by electronic mail to the coordinator center of the “Fistula Day” as proof of participation. Similarly, data collected in this first examination were locally entered into an on-line application built with RedCap®©^‡^ (University of Vanderbilt, United States).

The other two surveys contemplated in the “Fistula Day” project were completed at 30 days (June 9th 2018) and 60 days (July 9th, 2018) after the first one in order to record the evolution of the patient, and the therapeutic response achieved. No new cases were admitted in the subsequent “Fistula Day” examinations.

### Data processing and statistical-mathematical analysis of the results

R program for statistical management and analysis (R Core Team 2018 version 3.5.0, United States) was used for debugging, preparing and processing data collected during the “Fistula Day” surveys. Data were reduced to absolute | relative frequencies and percentages regarding variable type and the objective of statistical analysis.

Condition of the patient (Alive | Deceased), hospital stay (Prolonged | Discharged) and spontaneous closure (Yes | No) of the fistula were assumed as 30- and 60 days-outcomes of the “Fistula Day”. Nature and strength of the associations between the results of the “Fistula Day”, on one hand, and the characteristics of hospitals and patients included in the cohort, on the other, were examined with appropriate statistical tests regarding the variable type. Existent differences between cohorts of patients according with the selected predictor were evaluated with the log-rank test based on the chi-square distribution.^8^ A level lower than 5 % was used in all the instances for denoting the finding as significant.

### Treatment of missing data

Data lost during follow-up of the patient were replaced with the observation entered in the preceding examination according with the LOCF (for “*Last Observation Carried Forward*”) method.

### Intention-to-treat

Data collected during the “Fistula Day” were analyzed according with the “Intention to treat” principle in order to keep the size of the cohort constant in each of the surveys foreseen in the cohort.^9^

### Ethical considerations

The protocol followed by local surveyors during the “Fistula Day” was drafted according with “Good Clinical Practices”.^10^ Identity and rights of the surveyed patients were also protected.^11^ The patients (and by extension their caretakers) were informed about the purposes of the research, and the non-invasive nature of the procedures. Collected data were adequately preserved in order to ensure anonymity and confidentiality. Aggregated data were used in the interpretation of the results and the realization of statistical inferences. Informed consent was obtained from the patient before inclusion in the cohort. Local conduction of the activities foreseen in the “Fistula Day” was authorized and supervised by the hospital Committees of Ethics after presentation, review and approval of the research protocols.

The researchers entrusted with the conduction of the “Fistula Day Project” presented the protocol “Current status of the postoperative fistula of digestive tract; multicentric, multinational study. DAY OF THE FISTULA” before the Ethics Committee of the San Javier Hospital (city of Guadalajara, State of Jalisco, México) for review and approval. A ruling was emitted on April 11^th^, 2018 by Dr. Eduardo Razón Gutiérrez, acting Director of the Ethics Committee, with the approval of the research protocol and the authorization for the conduction of the “Fistula Day” Project^§^.

## RESULTS

At the conclusion of the first “Fistula Day” examination 177 patients diagnosed with, and treated for, GIF in 76 hospitals of LATAM (13 countries) and Europe (4) were admitted. The presence of patients attended in non-LATAM hospitals corresponded with the medical care teams’ desires to be included in the study cohort and sharing the locally collected data. However, non-LATAM patients amounted less than 10 % of the size of the cohort. On the other hand, half plus one of the patients admitted in the cohort were Mexicans.

### Influence of the demographical, clinical and sanitary characteristics of the patients upon prognosis and outcomes of GIF

Table 1 shows demographical, clinical and sanitary characteristics of the patients finally included in the “Fistula Day” study serie. Men prevailed over women (*Males*: 58.2 % of the study serie). Average age was 51.0 ± 16.7 years. Subjects with ages ≥ 60 years were 36.2 % of the studied cases. Fifty-nine-point-six percent of the patients accumulated between 0 – 30 days of hospital stay on admission into the study serie. A cancer diagnosis was made in 27.7 % of the patients.

**Table 1.**
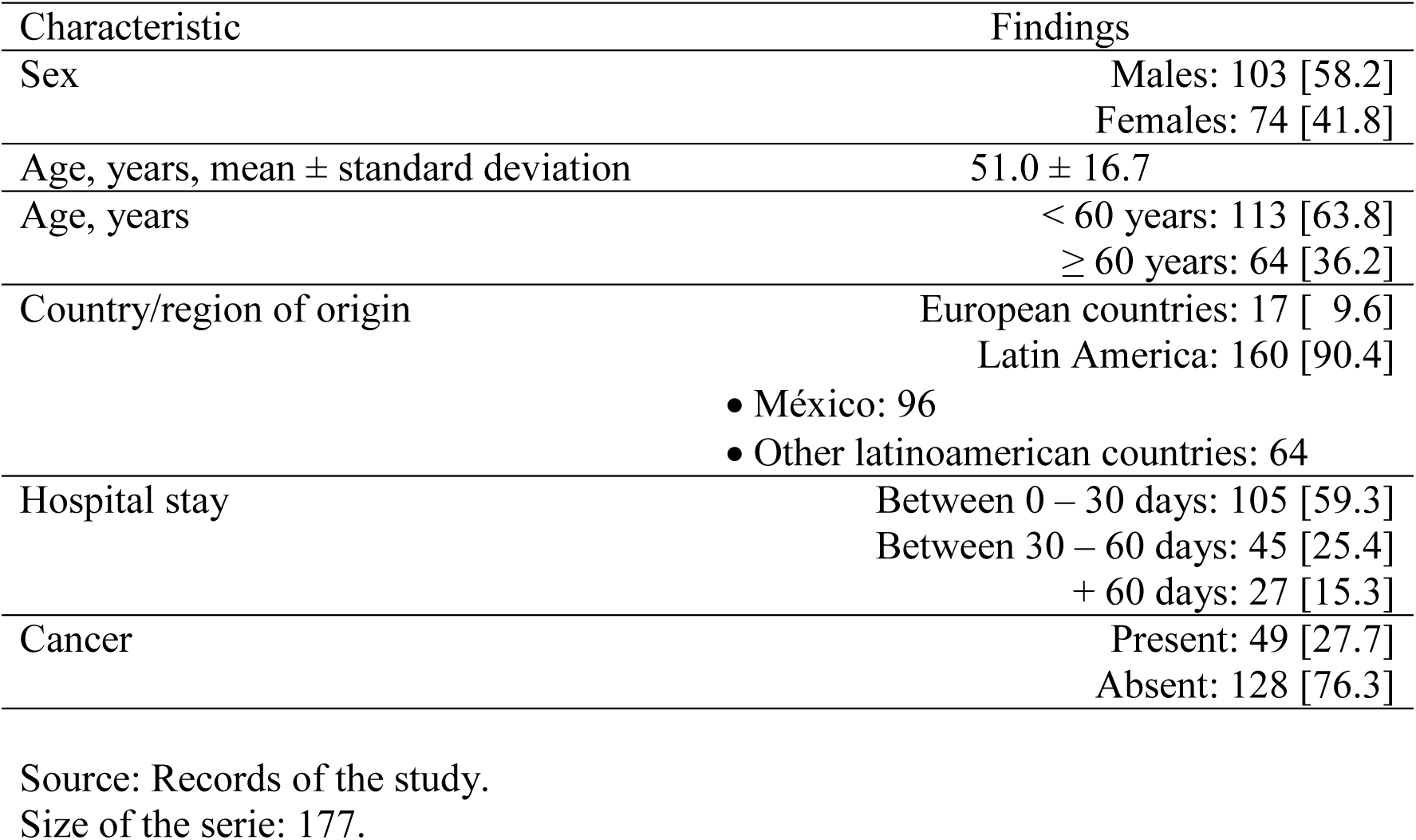
Demographical and clinical characteristics of the patients included in the cohort for studying the prognosis and outcomes of gastrointestinal fistulas. Number and [within bracketts] the percentage of the patients included in each category of the characteristic are shown. Mean ± standard deviation of the characteristic is shown in selected instance. For further details: See the text of the present essay.

| Characteristic | Findings |
| --- | --- |
| Sex | Males: 103 [58.2]<br>Females: 74 [41.8] |
| Age, years, mean ± standard deviation | 51.0 ± 16.7 |
| Age, years | < 60 years: 113 [63.8]<br>≥ 60 years: 64 [36.2] |
| Country/region of origin | European countries: 17 [ 9.6]<br>Latin America: 160 [90.4]<br>• México: 96<br>• Other latinoamerican countries: 64 |
| Hospital stay | Between 0 – 30 days: 105 [59.3]<br>Between 30 – 60 days: 45 [25.4]<br>+ 60 days: 27 [15.3] |
| Cancer | Present: 49 [27.7]<br>Absent: 128 [76.3] |

Source: Records of the study.
Size of the serie: 177.

On the closure of the “Fistula Day” window of observation, 26 deaths were registered (14.7 % of the size of the cohort) along with 84 prolonged hospitalizations (47.4 %). Spontaneous closure of the GIF was reported in 64 (36.1 %) of the examined patients. Table 2 shows the 30- and 60-days “Fistula Day” outcomes. Twenty patients (11.3 % of the size of the cohort) were lost to follow-up. In spite of this, estimates of the indicators of prognosis and outcomes of GIF were independent from the method of analysis used.

**Table 2.** Characteristics of the cohort of the study for studying the prognosis and outcomes of the gastrointestinal fistulas in the different moments of completion of the “Fistula Day” project. For further details: See the text of the present essay.

| Moment | 30 days follow-up |  | 60 days follow-up |  |
| --- | --- | --- | --- | --- |
| Method of analysis | *Intention-to-treat* | *Analysis-per-protocol* | *Intention-to-treat* | *Analysis-per-protocol* |
| Size of the serie |  |  |  |  |
| • Expected | 177 | 165 | 177 | 157 |
| • Lost to follow-up |  | 12 |  | 20 |
| Condition of the paient |  |  |  |  |
| • Alive | 158 [89.3] | 146 [88.5] | 151 [85.3] | 131 [83.4] |
| • Deceased | 19 [10.7] | 19 [11.5] | 26 [14.7] | 26 [16.6] |
| Hospitalized | 63<br>[35.6] | 63<br>[38.2] | 84<br>[47.4] | 84<br>[53.5] |
| Spontaneous closure | 60<br>[33.9] | 60<br>[36.4] | 64<br>[36.2] | 64<br>[40.8] |

Source: Records of the study.
Size of the serie: 177.

Table 3 shows the associations between demographical, clinical and sanitary characteristics of the patients and the indicators of the study’s outcomes. As shown, study’s results were independent from the demographical, clinical and sanitary characteristics of the patients.

**Table 3.** Associations observed between outcomes indicators of the study and the demographical, clinical and sanitary characteristics of the examined patients. Percentage of patients included in each stratum of the characteristic is shown in each instance. For further details: See the text of the present essay.

| Characteristic | Findings | Interpretation |
| --- | --- | --- |
| ***Survival of the patient*** |  |  |
| • Sex of the patient | Male: 87.4<br>Female: 82.4 | $\chi^2 = 0.841$ |
| • Age of the patient | < 60 years: 85.0<br>≥ 60 years: 85.9 | $\chi^2 = 0.031$ |
| • Diagnosis of cancer | Present: 87.8<br>Absent: 84.4 | $\chi^2 = 0.323$ |
| • Previous hospital stay | 0 – 30 days: 81.9<br>31 – 60 days: 94.3<br>> 60 days: 85.2 | $\chi^2 = 3.283$ |
| ***Prolonged hospitalization*** |  |  |
| • Sex of the patient | Male: 50.5<br>Female: 43.2 | $\chi^2 = 0.906$ |
| • Age of the patient | < 60 years: 52.2<br>≥ 60 years: 39.1 | $\chi^2 = 2.833$ |
| • Diagnosis of cancer | Present: 55.1<br>Absent: 44.5 | $\chi^2 = 1.538$ |
| • Previous hospital stay | 0 – 30 days: 50.5<br>31 – 60 days: 42.2<br>> 60 days: 44.4 | $\chi^2 = 0.977$ |
| ***Spontaneous closure of the fistula*** |  |  |
| • Sex of the patient | Masculino: 39.8<br>Femenino: 31.1 | $\chi^2 = 1.420$ |
| • Age of the patient | < 60 years: 34.5<br>≥ 60 years: 39.1 | $\chi^2 = 0.366$ |
| • Diagnosis of cancer | Present: 36.7<br>Absent: 35.9 | $\chi^2 = 0.009$ |
| • Previous hospital stay | 0 – 30 days: 34.3<br>31 – 60 days: 42.2<br>> 60 days: 33.3 | $\chi^2 = 0.969$ |

Source: Records of the study.
Size of the serie: 177.

Figures 1 – 3 show the behavior of the cohort as disaggregated regarding the demographical, clinical and sanitary characteristics of the studied patients in order to assess the prognosis and outcomes of GIF. As shown, behavior of the cohort in each case was independent from the demographical, clinical and sanitary characteristics of the studied patients.

**Figure 1.**
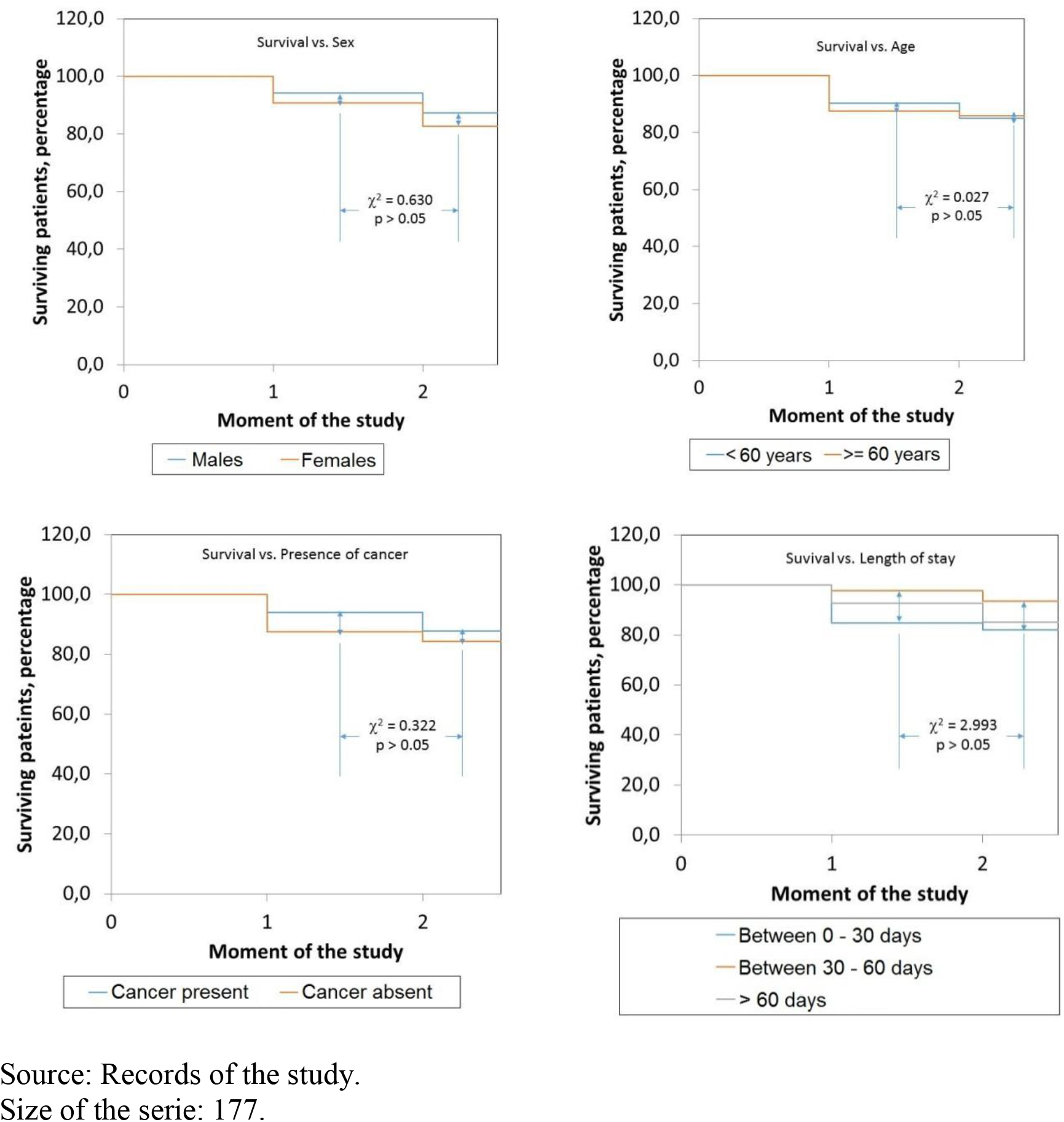
Behavior of the survival of the patients included in the cohort according with the demographical, clinical and sanitary characteristics as documented on admission in the “Fistula Day” project. For further details: See the text of the present essay.

**Figure 2.**
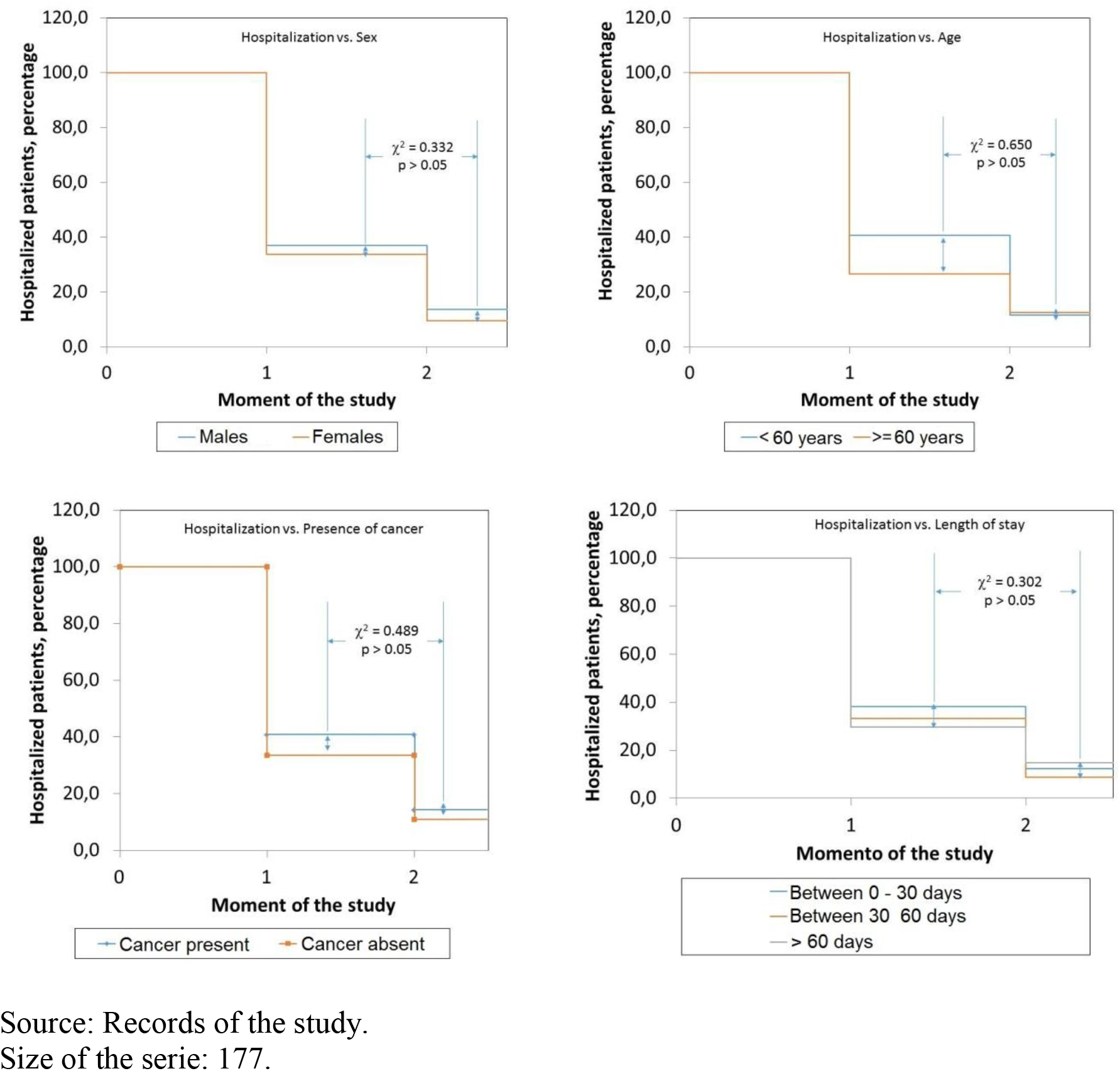
Behavior of the hospitalization of the patients included in the cohort according with the demographical, clinical and sanitary characteristics as documented on admission in the “Fistula Day” project. For further details: See the text of the present essay.

**Figure 3.**
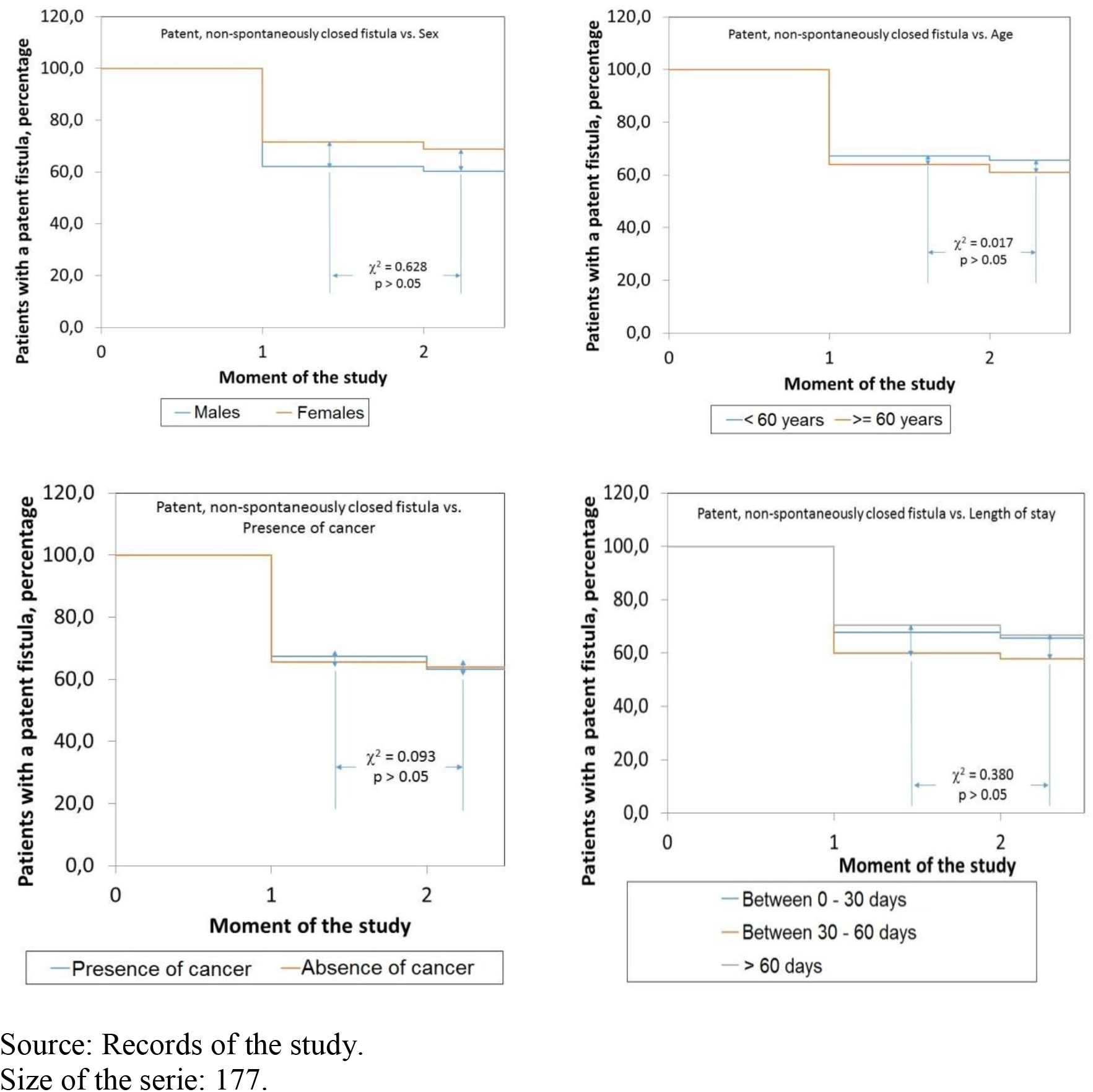
Behavior of the spontaneous closure of gastrointestinal fistulas in the patients included in the cohort according with the demographical, clinical and sanitary characteristics as documented on admission in the “Fistula Day” project. Curves were constructed with patients with a patent, non-spontaneously closed fistula, in the corresponding moment of the study. For further details: See the text of the present essay.

### Influence of the characteristics of the gastrointestinal fistulas upon prognosis and outcomes of the fistula

Table 4 shows the characteristics of the GIF documented in this study. Patients with ECF prevailed. Almost 60 % of the GIF showed an output < 500 mL/day. Small bowel and colon were the predominant locations as source of the GIF. Half plus one of the GIF was diagnosed after elapsing the first 5 days of the primary surgery. In addition, 60.5% of the GIF originated after an emergency surgery. It is to be noticed that a bariatric surgery was completed in less than 4 % of the studied patients.

**Table 4.** Characteristics of the gastrointestinal fistulas diagnosed in the patients participating in “Fistula Day” project. Number and [within bracketts] the percentage of the patients included in each category of the characteristic are shown. Mean ± standard deviation of the characteristic is shown in selected instance. For further details: See the text of the present essay.

| Characteristics of the fistulas | Findings |
| --- | --- |
| Fistulas diagnosed after | During the first 5 days after surgery: 84 [47.5]<br>5 days after surgery: 93 [52.5] |
| Type of primary surgery | Elective surgery: 70 [39.5]<br>Emergency surgery: 107 [60.5] |
| Nature of the fistula | Enterocutaneous fistulas: 115 [64.9]<br>Enteroathmosperic fistulas: 62 [35.1] |
| Location of the fistula | Esophagus: 12 [ 6.8]<br>Stomach: 18 [10.2]<br>Small bowel: 88 [49.7]<br>Colon and rectum: 49 [27.7]<br>Other locations/No declared: 10 [ 5.6] |
| Output of the fistula | Low: < 500 mL/24 hours: 106 [59.9]<br>High: ≥ 500 mL/24 hours: 40 [22.6]<br>No output/No declared: 31 [17.5] |

Source: Records of the study. Size of the serie: 177.

Table 5 shows the associations between GIF characteristics and the outcomes indicators of the study. Type of GIF influenced upon 60-days survival (and mortality as a complementary finding) of the patient: survival of the patient was lower among patients with an EAF: *EAF*: 82.3 % vs. *ECF*: 87.0 % (Δ = −4.7 %; *χ*^2^ = 6.878; p < 0.05). Type of surgery previously performed also influenced upon prolongation of hospital stay: *Elective surgery*: 61.4 % vs. *Emergency surgery*: 38.3 % (Δ = +23.1 %; *χ*^2^ = 9.064; p < 0.05).

**Table 5.**
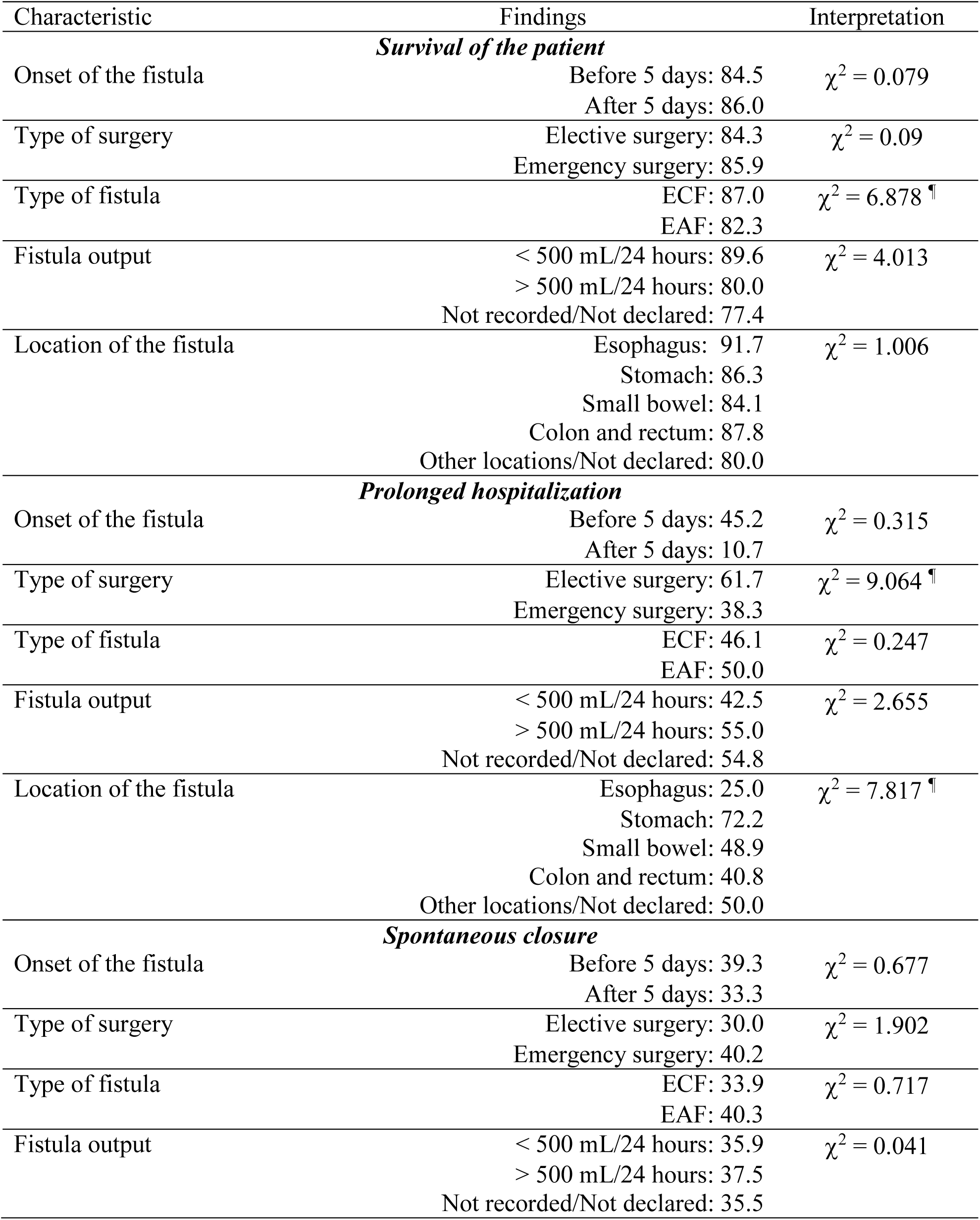
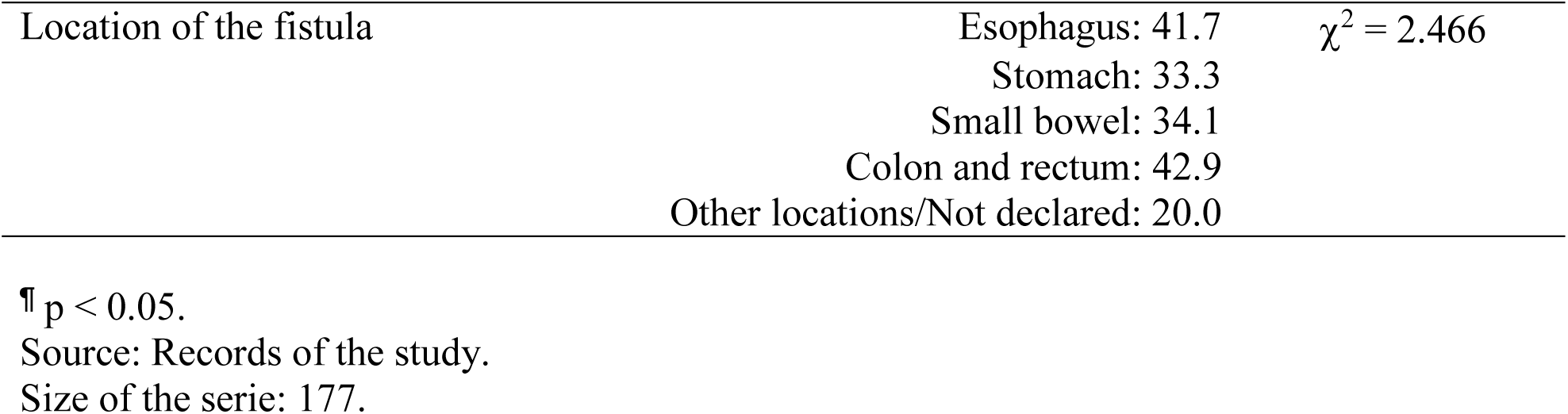
Associations observed the outcomes indicators of the study and the characteristics of the gastrointestinal fistulas. Percentage of patients included in each stratum of the characteristic is shown in each instance. For further details: See the text of the present essay.

Similarly, influence of the location of the fistula upon prolongation of the hospitalization is to be signaled: patients with a fistula originated in the stomach (72.2 % of those within this location), small bowel (48.9 %), and colon and rectum (48.1 %) exhibited the highest rates of prolonged hospitalization (*χ*^2^ = 7.817; p < 0.05). However, uneven distribution of the different locations of GIF should not be overlooked; a common finding in observational studies.

Figures 4 – 6 show the influence of the GIF characteristics upon the behavior of the cohort. Only the type of GIF influenced upon survival of the patient: the number of surviving subjects was always among those with an ECF: *At 30 days*: ECF: 92.1 % vs. EAF: 83.9 % (Δ = +8.2 %); *At 60 days*: ECF: 98.1 % vs. EAF: 90.4 % (Δ = +7.7 %; *χ*^2^ = 13.764; p < 0.05). On the contrary, cohorts of patients disaggregated regarding type of previous surgery did not differ between them: *At 30 days*: Elective surgery: 44.3 % vs. Emergency surgery: 29.9 % (Δ = +14.4 %); *At 60 days*: Elective surgery: 30.8 % vs. Emergency surgery: 12.0 % (Δ = +18.8 %; *χ*^2^ = 2.975; p = 0.0845). Cohorts of patients disaggregated according with location of fistula did not differ between them either (*χ*^2^ = 3.042; p > 0.05).

**Figure 4.**
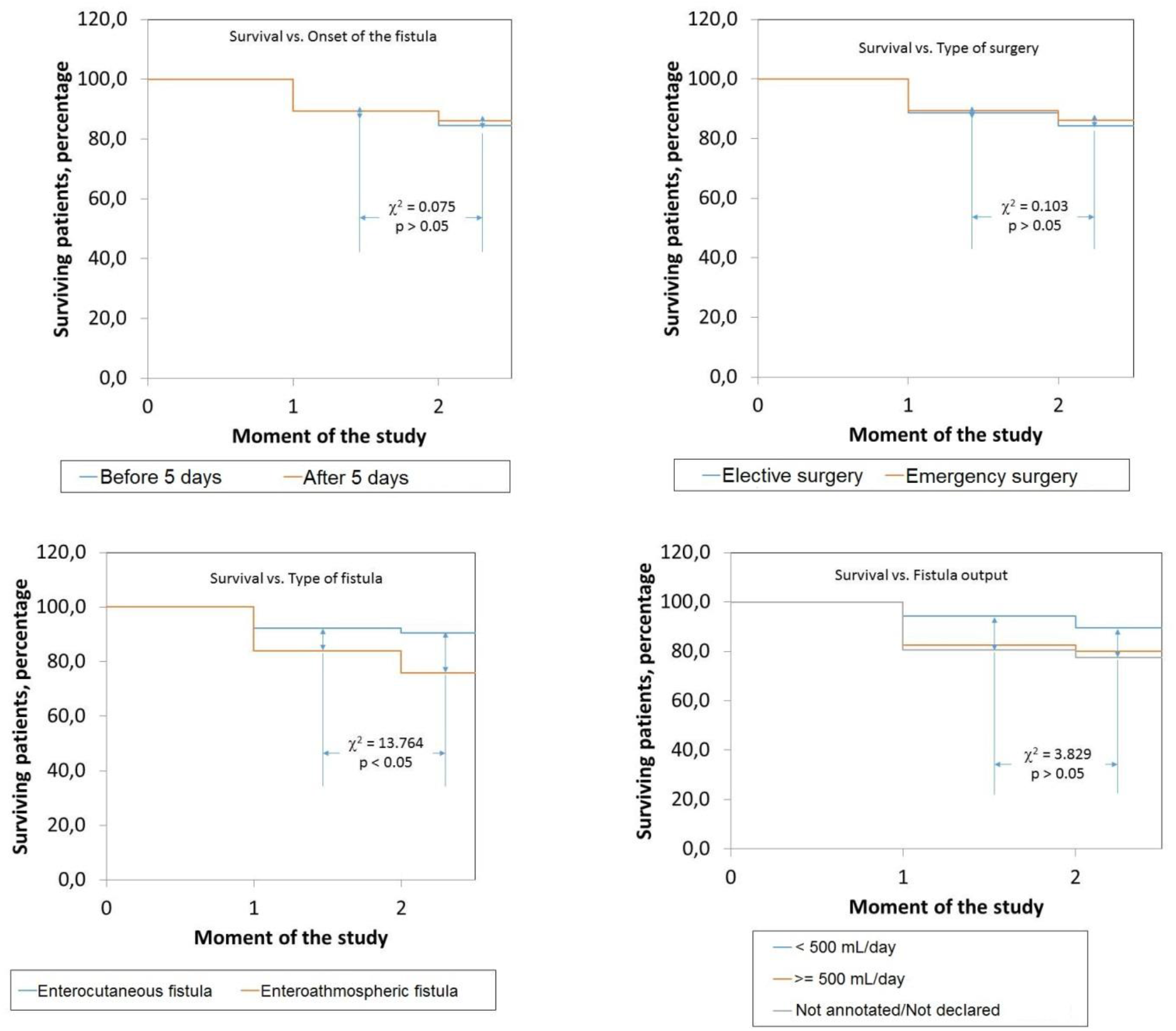
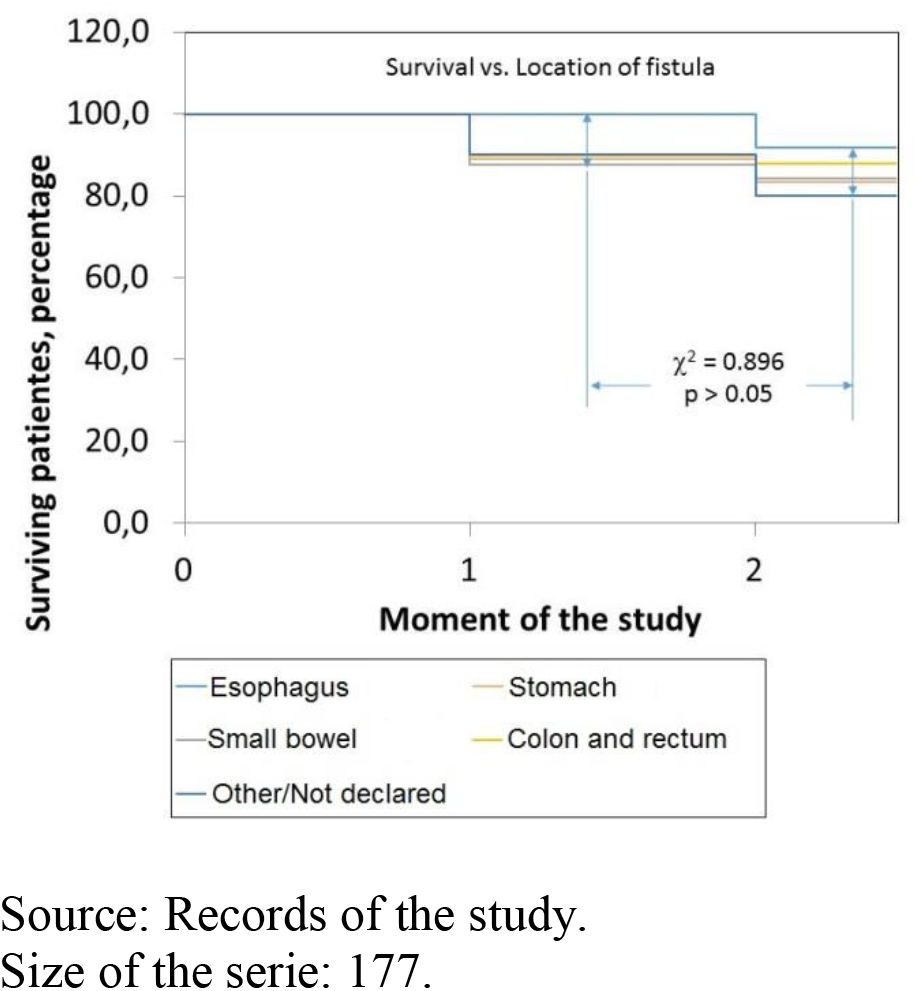
Behavior of the survival of the patient according with the characteristics of the gastrointestinal fistulas as documented on admission in the “Fistula Day” project. Influence of the type of the fistula upon accumulated survival is to be noticed. For further details: See the text of the present essay.

**Figure 5.**
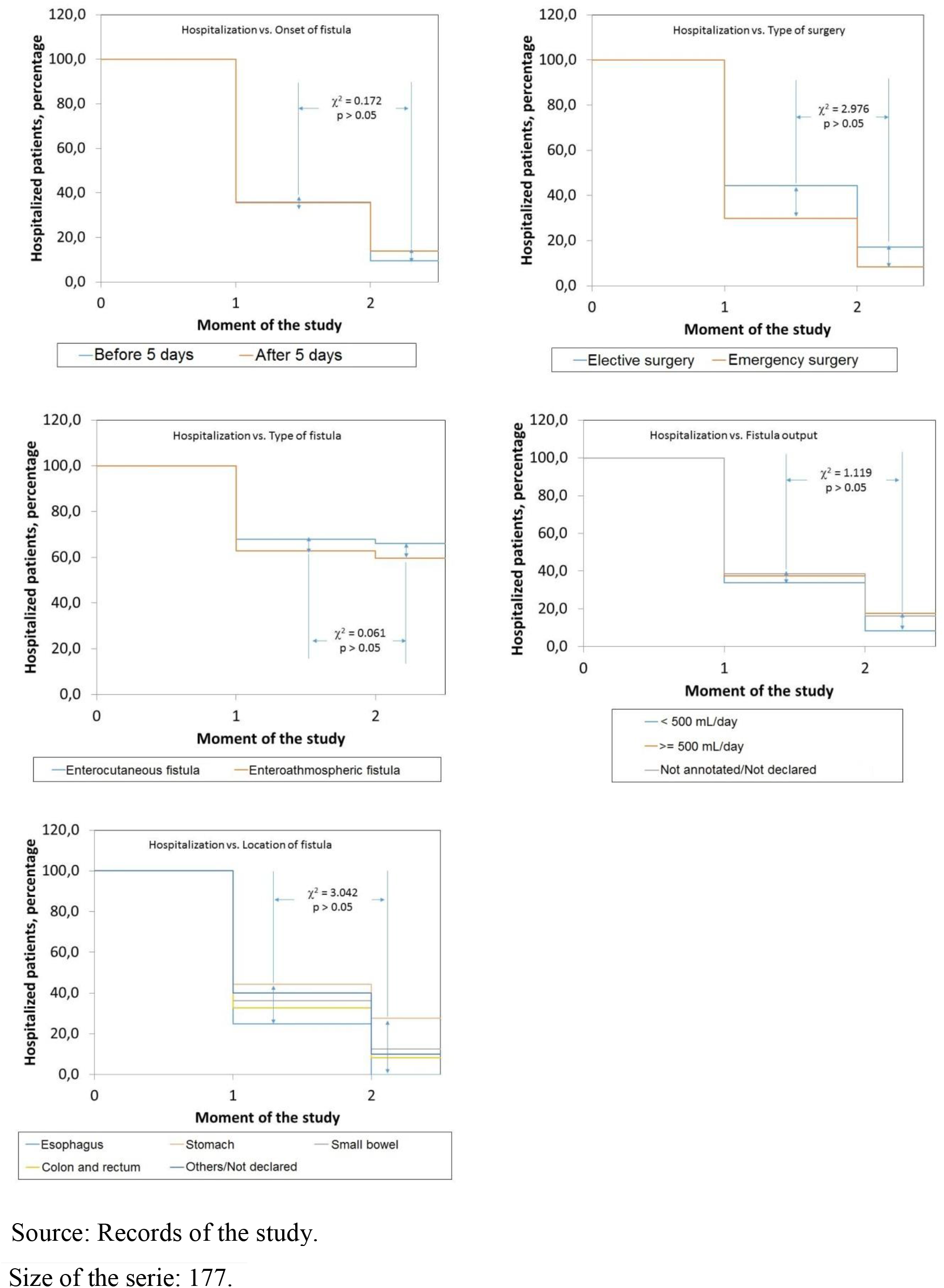
Behavior of the hospitalization of the patient according with the characteristics of gastrointestinal fistulas as documented on admission in the “Fistula Day” project. For further details: See the text of the present essay.

**Figure 6.**
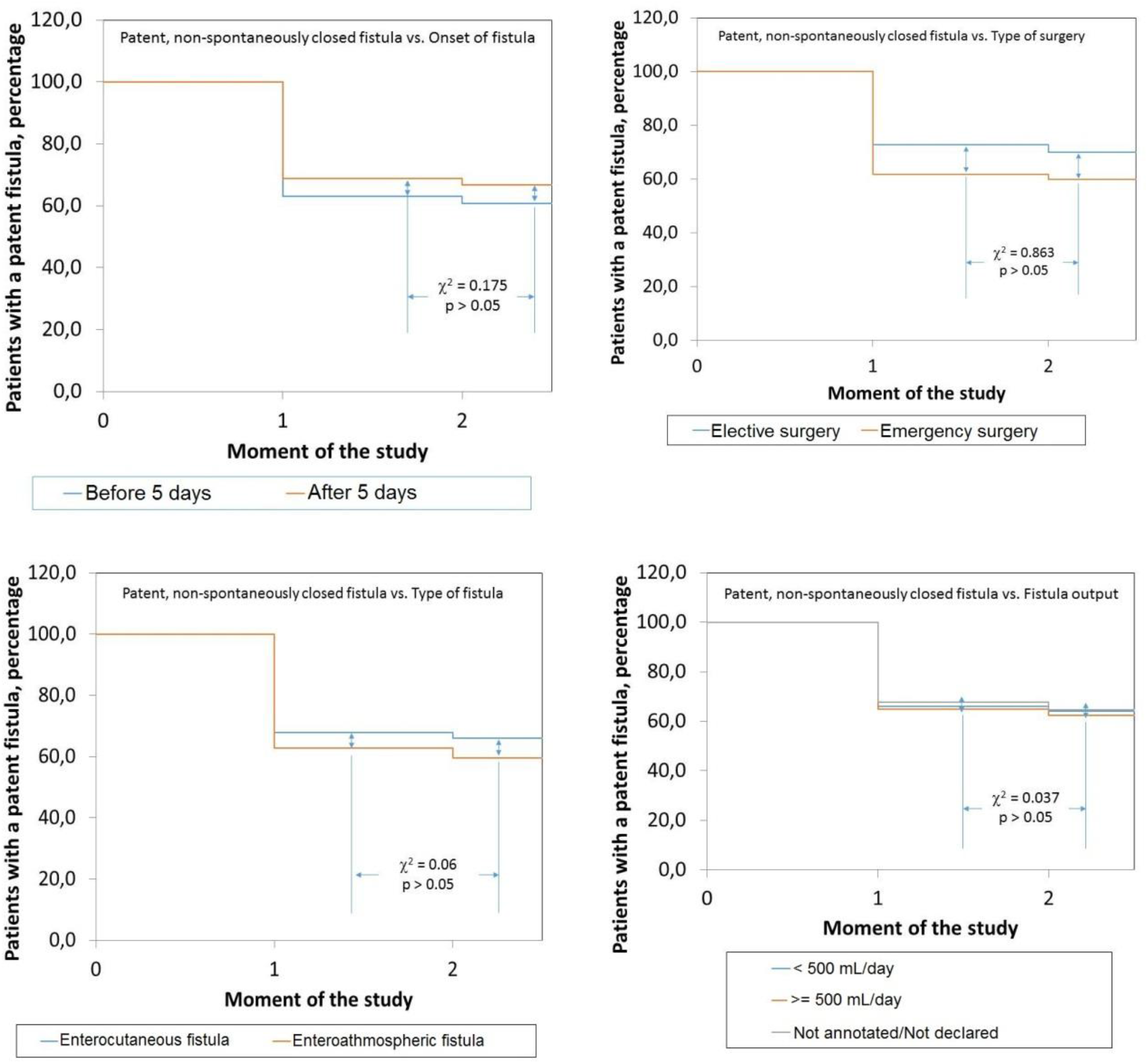
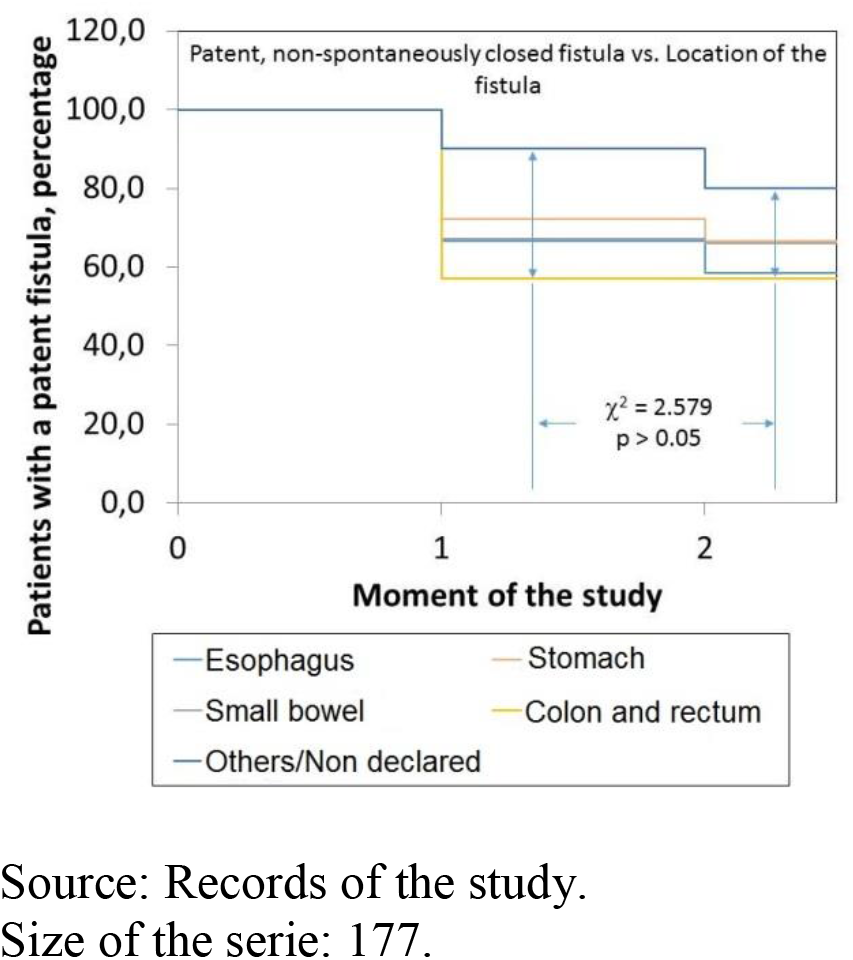
Behavior of the spontaneous closure of gastrointestinal fistulas in the patients included in the cohort according with the characteristics of the fistulas as documented on admission in the “Fistula Day” project. Curves were constructed with patients with a patent, non-spontaneously closed fistula, in the corresponding moment of the study. For further details: See the text of the present essay.

### Influence of the nutritional status of the patient

Table 6 shows the nutritional characteristics of the GIF patients. Average BMI was 24.3 ± 6.1 kg.m^-2^. Excessive body weight presented in 40.1 % of the cases. However, more than a third of the GIF patients referred weight loss > 10 % on admission in the study. Average calf circumference (CC) was 30.5 ± 16.9 cm. Half plus one of the patients presented with a CC < 31 cm: cut-off point used in the description of this indicator. It is to be noticed that CC was not measured in the tenth of the GIF patients. More than half of the GIF patients had a serum albumin < 30 g.L^-1^. Only 52.5 % of the C reactive protein (PCR) determinations were recovered: the reason foe excluding this nutritional indicator from the analyses.

**Table 6.** Nutritional characteristics of the patients participating in the “Fistula Day” project. Number and [within brackets] percentage of the patients included in each category of the characteristic are shown. Mean ± standard deviation of the characteristic are shown in selected instances. For further details: See the text of the present essay.

| Characteristic | Findings |
| --- | --- |
| Height, cm | 164.6 ± 9.3 |
| Body weight, kg | 66.0 ± 17.6 |
| Body Mass Index, kg.m<sup>-2</sup> | 24.3 ± 6.1 |
| Body Mass Index, kg.m<sup>-2</sup> | < 18.5: 26 [14.7]<br>18.5 – 24.9: 80 [45.2]<br>≥ 25.0: 71 [40.1] |
| Calf circumference, cm | 30.5 ± 16.9 |
| Calf circumference, cm | < 31.0: 98 [55.4]<br>≥ 31.0: 59 [31.1]<br>Not declared: 20 [11.3] |
| Weight loss, % | < 5.0 %: 66 [37.3]<br>5 – 10 %: 44 [24.9]<br>10 – 15 %: 36 [20.3]<br>> 20 %: 31 [17.5] |
| Serum albumin, g.L<sup>-1</sup> | ≤ 30: 103 [58.2]<br>> 30: 64 [36.1]<br>Not declared: 10 [ 5.6] |
| C reactive protein, mg.L<sup>-1</sup> | < 0.56: 7 [ 3.9]<br>0.56 – 4.00: 0 [ 0.0]<br>5.00 – 50.00: 3 [ 1.7]<br>51.00 – 100.00: 4 [ 2.3]<br>101.00 – 200.00: 18 [10.2]<br>> 200.00: 32 [18.1]<br>Not declared: 93 [52.5] |

Source: Records of the study. Size of the serie: 177.

Table 7 shows the associations between the indicators used to describe the nutritional status of GIF patient and outcomes indicators of the study. CC influenced upon prolongation of hospital stay of GIF patients: patients with a diminished CC showed a more prolonged hospital stay: *CC < 31 cm*: 60.3 %; *CC ≥ 31 cm*: 39.0 %; *CC not annotated/not declared*: 26.7 % (*χ*^2^ = 12.655; p < 0.05; independence test based on the chi-squared distribution). On the other hand, weight loss experienced by the patient exerted a marginal effect upon prolongation of hospital stay (*χ*^2^ = 7.294; p = 0.063).

**Table 7.** Associations observed within the indicators of the nutritional status of the patients with gastrointestinal fistulas and the indicators of prognosis and outcomes of the patient. Percentage of patients included in each stratum of the category is shown in each instance. For further details: See the text of the present essay.

| Characteristic | Findings | Interpretation |
| --- | --- | --- |
| ***Survival of the patient*** |  |  |
| Body Mass Index | < 18.5: 92.3<br>18.5 – 24.9: 81.3<br>≥ 25.0: 87.3 | $\chi^2 = 0.090$ |
| Calf circumference | < 31.0: 86.7<br>≥ 31.0: 84.7<br>Not declared: 80.0 | $\chi^2 = 0.623$ |
| Weight loss | < 5.0 %: 81.8<br>5 – 10 %: 86.4<br>10 – 15 %: 88.9<br>> 20 %: 87.1 | $\chi^2 = 1.128$ |
| Serum albumin, g.L<sup>-1</sup> | ≤ 30: 85.6<br>> 30: 85.9<br>Not declared: 80.0 | $\chi^2 = 0.246$ |
| ***Prolonged hospitalization*** |  |  |
| Body Mass Index | < 18.5: 30.8<br>18.5 – 24.9: 51.3<br>≥ 25.0: 49.3 | $\chi^2 = 3.461$ |
| Calf circumference | < 31.0: 60.3<br>≥ 31.0: 39.0<br>Not declared: 26.7 | $\chi^2 = 12.655$ ¶ |
| Weight loss | < 5.0 %: 50.0<br>5 – 10 %: 54.5<br>10 – 15 %: 52.8<br>> 20 %: 25.8 | $\chi^2 = 7.294$<br>p = 0.063 |
| Serum albumin, g.L<sup>-1</sup> | ≤ 30: 47.6<br>> 30: 43.7<br>Not declare: 70.0 | $\chi^2 = 2.391$ |
| ***Spontaneous closure*** |  |  |
| Body Mass Index | < 18.5: 42.3<br>18.5 – 24.9: 36.3<br>≥ 25.0: 33.8 | $\chi^2 = 0.09$ |
| Calf circumference | < 31.0: 33.0<br>≥ 31.0: 35.6<br>No declara: 40.0 | $\chi^2 = 0.144$ |
| Weight loss | < 5.0 %: 30.3<br>5 – 10 %: 38.6<br>10 – 15 %: 44.4<br>> 20 %: 35.5 | $\chi^2 = 2.174$ |
| Serum albumin, g.L<sup>-1</sup> | ≤ 3.0: 33.0 | $\chi^2 = 1.059$ |
|  | > 3.0: 40.6 |  |
|  | Not declared: 40.0 |  |

Source: Records of the study. Size of the serie: 177.

Lastly, Figures 7 – 9 show the influence of the nutritional indicators upon prognosis and outcomes of GIF. As it can be seen, nutritional status did not influence upon behavior of the study cohort regarding the indicators of prognosis and outcomes of GIF.

**Figure 7.**
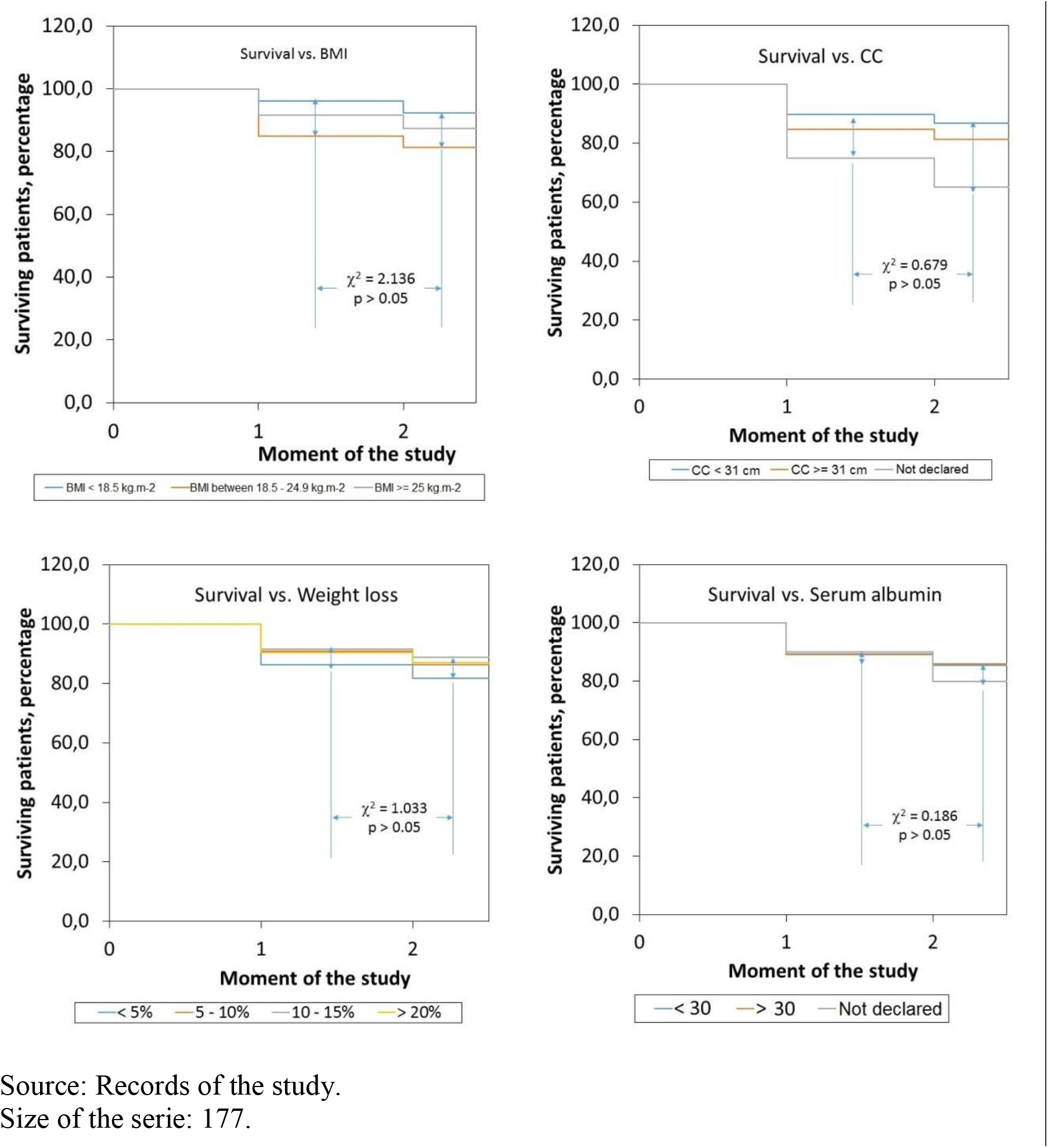
Behavior of the survival of the patient according with the nutritional characteristics of the patient as documented on admission in the “Fistula Day” project. For further details: See the text of the present essay. Legend: BMI: Body Mass Index. CC: Calf circumference.

**Figure 8.**
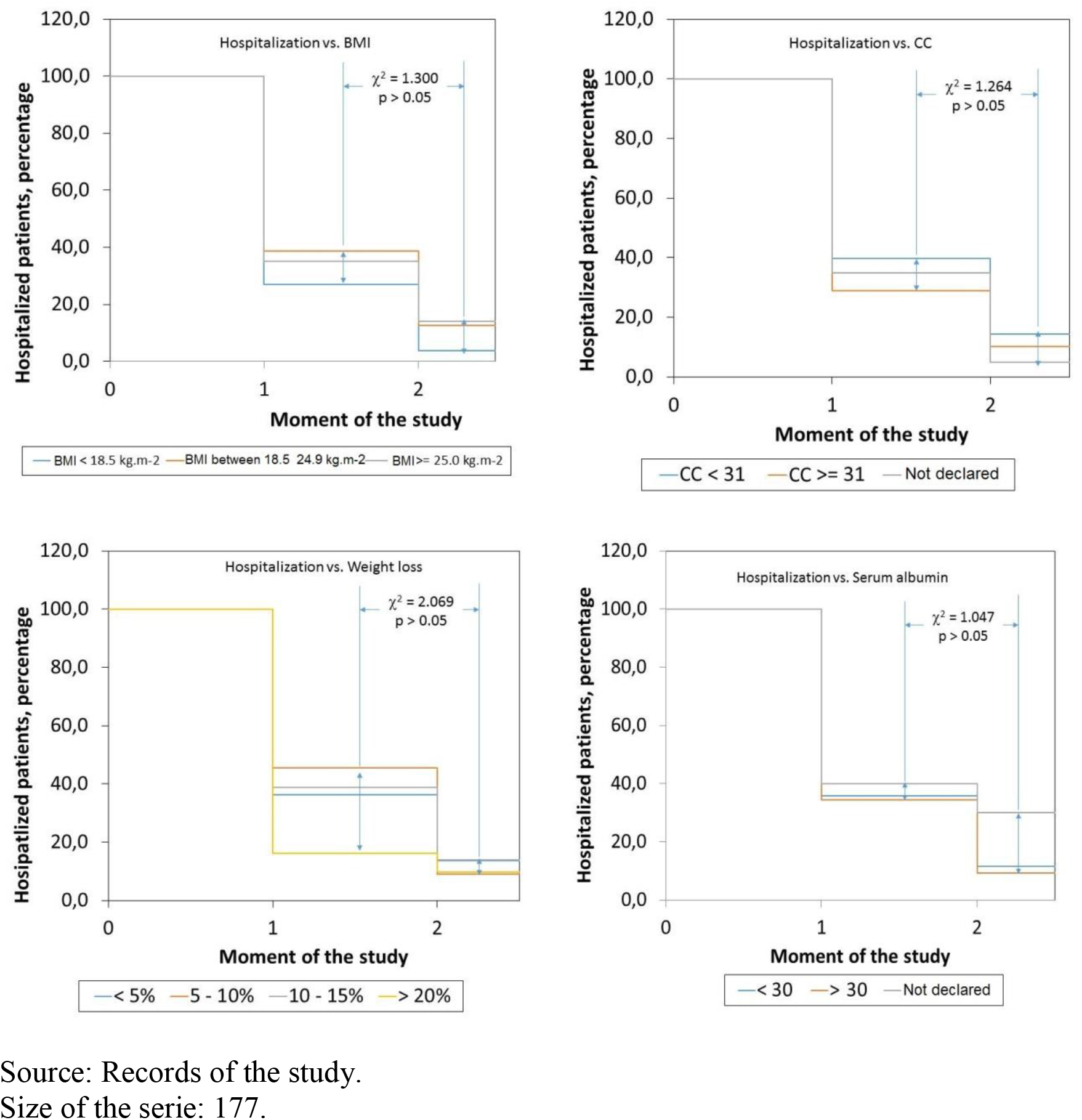
Behavior of the hospitalization according with the nutritional characteristics of the patient as documented on admission in the “Fistula Day” project. For further details: See the text of the present essay. Legend: BMI: Body Mass Index. CC: Calf circumference.

**Figura 9.**
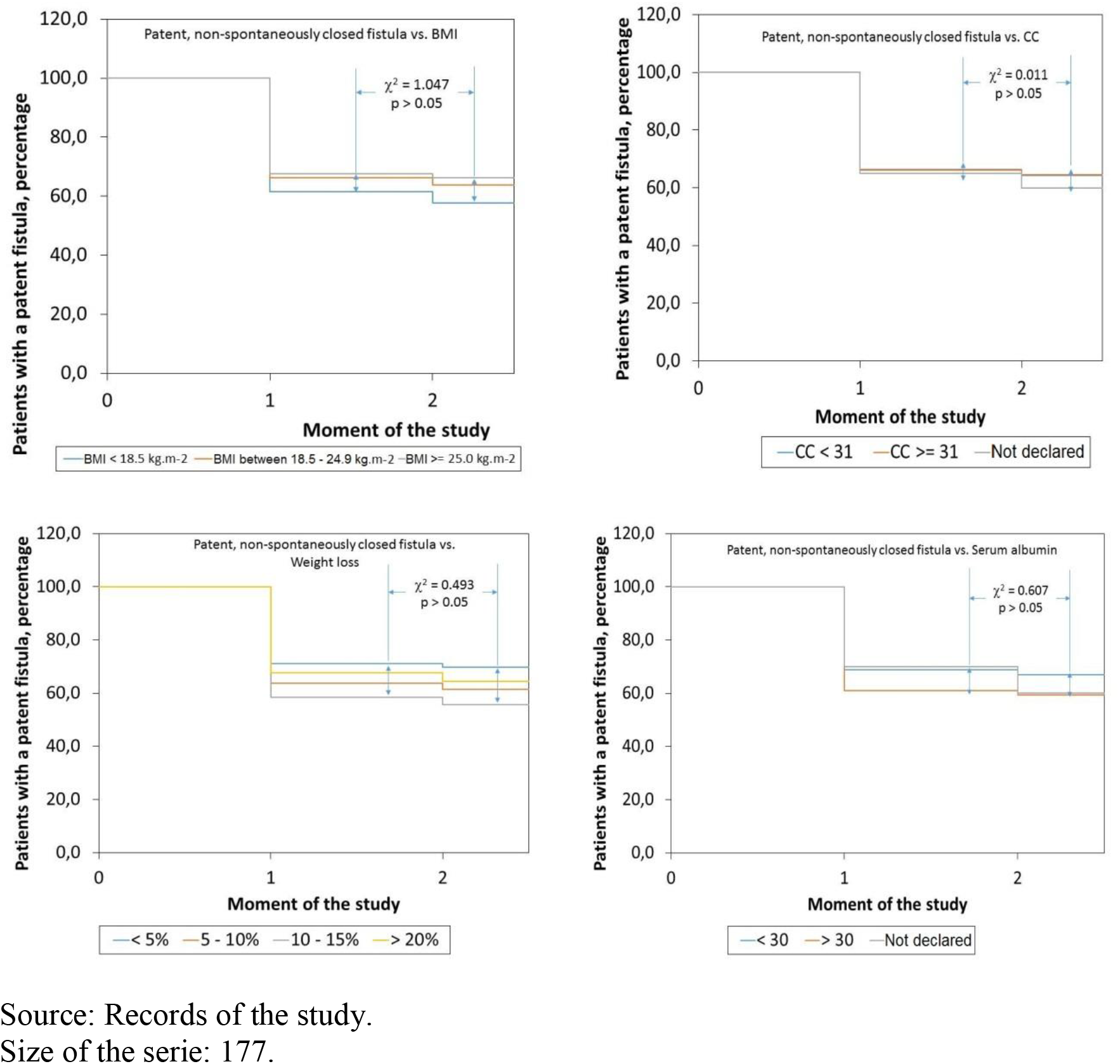
Behavior of the spontaneous closure of gastrointestinal fistulas in the patients included in the cohort according with their nutritional characteristics as documented on admission in the “Fistula Day” project. Curves were constructed with patients with a patent, non-spontaneously closed fistula, in the corresponding moment of the study. For further details: See the text of the present essay. Legend: BMI: Body Mass Index. CC: Calf circumference.

## DISCUSSION

The “Fistula Day” has been conceived as a multi-national, multi-center project aimed to reveal the practices currently followed in the diagnosis and treatment of GIF so, in this way, build the required evidences for the continuous improvement of the quality of surgical and clinical processes. Originally proposed as a Latin-American effort, the “Fistula Day” has also incorporated data provided by medical care teams from hospitals of the United Kingdom and the European Union that requested (and were accepted) to participate in this effort. In spite of this, “Fistula Day” might represent the current state of the treatment of GIF in Latin America, given the fact the majority of the patients being finally included in the database of the project was attended in Latin-American hospitals. It should also be pointed out that more than half of the studied patients were Mexicans: an expected finding giving the preponderance of Mexican health institutions in the database of the study.

The challenge in a research effort of such nature as the one described herein is to integrate all the collected data within a coherent model for interpreting the current reality of the treatment of the GIF. This essay attempted to assess how the characteristics of the patient, on one hand, and those of the fistula *per se*, on the other; influence upon the prognosis and the outcomes of the GFI through 3 indicators: survival of the patient, prolongation of the hospital stay, and the likely spontaneous closure of the fistula.

Most of the patients included in the “Fistula Day” database were men in their sixth decade of life accumulating between 0 – 30 days of hospital stay before admission in the present study. A third part of the patients had ages ≥ 60 years: a finding congruent with demographical aging and an increasingly presence of elderlies in the hospital populations. It is to be noticed that cancer was present as primary diagnosis in almost a third of the patients.

After conclusion of the first edition of the “Fistula Day” a 14.7 % mortality rate was observed. A systematic review with meta-analysis found a 3.0 % average mortality associated with enteral fistulas.^12^ Mortality could be as high as 7.0 % in patients awaiting for a GIF repairing surgery.^12^ The authors of the present essay also found a report mentioning a 20.0 % mortality after repair of enterocutaneous fistulas.^13^ On the other hand, Campos *et al*. (1999)^14^ reported a 30.9 % mortality rate in 188 patients diagnosed and treated with digestive fistulas (with biliopancreatic fistulas amounting for a fifth of them). Within this context, a 14.7 % mortality rate could be perceived as disproportionate, and would lead to further dwell in the causes for it, such as the characteristics of the patient, the fistula, and the hospital admitting and treating him/her, and its organization and the procedures conducted within the institution.

The aforementioned could be also applied to the other two indicators of the prognosis and outcomes of GIF. Hospital stay was prolonged in almost half of the surveyed patients. More than a tenth of the patients initially admitted in the database remained hospitalized after 60 days. Hospital resource is becoming increasingly costly in a setting marked by budgetary and fiscal cuts, along with a greater demand for services and positive impacts.^15^ Hence, even a rate > 10.0 % in the prolongation of hospitalization would imply additional economical burdens thus reducing opportunities for other equally needing patients.

Regarding the spontaneous closure of GIF (as the first option for containment and treatment), de Vries *et al*. (2017)^12^ have estimated a > 80.0 % total fistula closure rate reflecting the sums of actions taken in that direction (conservative as well as surgical ones). In the present study, spontaneous closure was achieved only in merely a third of the patients initially admitted in the study. A fistula closure (at least spontaneous) rate < 50.0 % might point towards failures, insufficiencies and inexperiences in GIF treatment. It is to be noticed Campos *et al*. (1999)^14^ reported in their study a 31.4 % spontaneous closure rate, when the international literature showed in that particular moment estimates as varied as 23.0 % and 80.0 %.^14^

It could be anticipated the characteristics of the patient might influence in the prognosis and outcomes of GIF, among them subject’s age, previous hospital stay, and cancer diagnosis. Hence, elderlies, patients with prolonged previous hospital stays, and those treated for cancer would distinguish themselves for a lower survival, prolonged hospitalization on closure of the observation window, and a lower rate of spontaneous closure. This was not the case: none of the characteristics of the patient decisively influenced upon the prognosis and outcomes of the fistula.

Similarly, and in correspondence with the accumulated literature, nutritional status of the GIF patient might determine the prognosis and outcomes of the fistula. Fistula is a catastrophic condition for all the domains of the economy, nutritional status in particular, and the resulting malnutrition, in turn, affects survival, prolongs hospital stay, and interferes with the spontaneous closure of the fistula; thus creating a vicious circle hard to break. In the present study, notwithstanding the fact subjects with a BMI ≥ 18.5 kg.m^-2^ prevailed (suggesting at first a preserved nutritional status), and excessive body weight presented in 40.1 % of the cases, more than a third of the patients referred > 10 % weight loss, half plus one of the patients presented with a diminished CC, and more than half of the GIF patients had a serum albumin < 30 g.L^-1^: these three indicators pointing to an important protein depletion in the study serie. Although protein depletion secondary to GIF could increase the mortality risk of the patient, prolong the hospital stay and/or extend the patency of the fistula, in the present study nutritional status did not influence either upon condition of the patient on the closure of the observation window nor the likelihood of spontaneous closure of the fistula. However, only reduced CC on admission in the study determined a prolonged hospital stay.

Influence of the characteristics of the GIF in its own on the outcomes indicators proposed in this study remains to be considered. It is to be noticed patients with an ECF prevailed, and in most of the instances fistula output was < 500 mL/day. It is also to be noticed most of the GIF originated after an emergency surgery. Regarding specialized reports declaring a high frequency of GIF after completion of a bariatric surgery,^16-17^ rate of conduction of these procedures was < 4.0 % in this study serie. Observation of small bowel and colon as the predominant locations of GIF is in line with consulted reports.^12-14^ It was then interesting to see that type of GIF determined subject’s survival, and (as a complementary phenomenon) the higher mortality observed in this study concentrated among patients with an EAF. Exposure of the mucosa has been cited in the specialized literature as one of the factors attempting against the spontaneous closure of the fistula, thus translating to an increased risk of complications (death among them).^18^

Evolution and outcomes of GIF might also be dependent upon location of fistula and the type of surgery associated with this complication, at least, when it comes to prolongation of hospitalization of the patient. Locations of the fistula associated with different rates of prolonged hospitalizations, with fistulas originated in the stomach being associated with the highest rate. However, data paucity (given by the uneven distribution of the cases within the different strata of this category, as it is the case with observational studies) might have obscure this finding. On the other hand, the type of surgery associated with the origin of the fistula might also lead to longer hospitalization, with ECF patients consuming prolonged hospital stays probably because of the conduction of an augmented number of procedures to achieve closure of the fistula.

## CONCLUSIONS

Evolution and outcomes of GIF might be dependent mainly upon type of the fistula. This association might be “colored” by the location of the fistula and the type of surgery linked with the origin of the fistula, these characteristics determining prolongation of hospital stay. Protein depletion secondary to GIF might also prolong hospital stay of the patient

### Future extensions

Influence of the characteristics of the patient and of the fistula in the evolution and outcomes of GIF has been examined in this essay. However, a comprehensive analysis of this issue would demand the exam of the influence of the characteristics of the hospital. In addition, the methodology of analysis discussed herein would also allow to assess how medical care teams face and deal locally with GIF. These questions will be satisfied in consecutive deliveries.

## Supporting information

Strobe check-list for the article

Letter of Approval by the San Javier Hospital Ethics Committee

## Data Availability

Interested researchers should address the authors for policies regarding access to the study data.

## ANNEXES

Annex 1. Countries and surveyors participating in the “Fistula Day” project.

***México***: Alejandro Serrano, Ali Navarrete Salgado, Aniriam García, Carla Beylan O‘Horan, Carlos Adolfo Andrade Ortega, Carlos Solar Aguirre, César Cruz Lozano, César Romero Mejía, Daniel de la Cruz Álvarez, Daniel Guerra Melgar, Emilio Prieto Díaz Chávez, Ernesto Duarte, Francisco García Morales, Gerardo de la Torre, Gilberto Ariel Arévalos Gómez, Gustavo Ervet Arias Quiñones, Heliodoro Plata Álvarez Area, Humberto Arenas Márquez, Javier Carrillo, Javier Luna Martínez, Javier Sánchez, Jorge Arturo Leal Martínez, Jorge Fernández Álvarez, José Alberto Carbajal, José Roberto Ramírez Espejo, Juan Antonio Delgado, Juan Carlos Hernández Aranda, Juan Carlos Salinas Vigueras, Julio Naranjo, Karla Leohner Ruezga, Lizeth Castillo Domínguez, Luciano Héctor Rogel Ríos, Luis Ibarra, Luis Ricardo Ramírez, Mabel Serrano, María del Carmen Aburto Fernández, Miguel Rivera, Mónica Patricia Bejarano Rosales, Ramón Mariscal Juárez Islas, Raúl Hernández Centeno, Rey Romero, Rogelio Galeno, Rogelio Ulises Pérez Vázquez, Salvador Almanza; ***Argentina***: Daniel Wainstein; ***Bolivia***: Ricardo Pérez Aliaga, Luis Bustamante, Milenka Aguilar Calle; ***Brasil***: José Aguilar do Nascimiento, Isabel Correia, José Henrique Agner Riveiro, Karoline Calfa Pitanga; ***Colombia***: Arturo Vergara, Charles Bermúdez, Angela Navas; ***Cuba***: Sergio Santana, Aldo Álvarez, David León, Jesús Barreto, Luis Garcés García-Espinosa, Julia Pupo, Antúan Quintero; ***Ecuador***: Dolores Rodríguez Veintimilla, María Eloisa García Velásquez, Alex Vasconez, Dolores Jima; ***Panamá***: Alfredo Matos; ***Paraguay***: Laura Joy; ***Perú***: Noemi Matilde, Eduardo Huaman, Mario Ferreira, Néstor Palacios, Melina Duffo; ***Uruguay***: Santiago Pose, Eduardo Moreira, Carlos Barozzi; ***España***: José María Jover, Patricia Villaroel, María Maiz, Loredana Arhip; ***Francia***: Francisca Joly, Stephane Schneider; ***Inglaterra***: Alastair Forbes; ***Polonia***: Stanislaw Klek.

## Footnotes

* From heretofore regarded as FELANPE.

† Disponible en: http://www.Fistuladay.org.

‡ Disponible en: http://www.redcap.org.

§ A photocopy of the ruling by the Ethics Committee of the San Javier Hospital is included as a supplemental material.

