## Supplementary material for "Outcomes of gastrointestinal fistulas: Results from a multi-continent, multi-national, multi-center cohort": Strobe check-list for the article

Humberto Arenas Márquez. MD, FACS

María Isabel Turcios Correia. MD, PhD

Juan Francisco García. MD, FACS

Roberto Anaya Prado. MD, FACS

Arturo Vergara. MD, FACS

Jorge Luis Garnica. MD

Alejandra Cacho. MD

Daniel Guerra. MD

Miguel Mendoza. MD

Sergio Santana Porbén. MD

**Affiliation**:

Unidad de Falla intestinal SANVITE

Hospital San Javier

Avenida Pablo Casals 640

Prados Providencia

Guadalajara 44670

Estado de Jalisco

Estados Unidos Mexicanos

Address all correspondence to:

Humberto Arenas Márquez

Electronic address:

**SUMMARY**

***Rationale***: Gastrointestinal fistulas (GIF) represent a severe and potentially lethal complication in hospital surgical patients. However, evidences are lacking about prognosis and outcomes of GIF in Latin America (LATAM) hospitals. ***Objective***: To describe the prognosis and outcomes GIF in LATAM hospitals. ***Study design***: Prospective, longitudinal, cohort-type study. The cohort fostered three cross-sectional examinations: First examination: On admission of the patient in the study; Second examination: Thirty days later; and Third (and last) examination: Sixty days after patient´s admission. ***Study serie***: One hundred seventy-seven patients (*Males*: 58.2 %; *Average age*: 51.0 ± 16.7 years; Ages ≥ 60 years: 36.2 %) diagnosed with, and assisted for, GIF (ECF: Enterocutaneous: 64.9 % *vs*. EAF: Enteroathmospheric: 35.1 %) in 76 LATAM hospitals (13 countries) and Europe (4). ***Methods***: Condition (Alive *vs*. Deceased) and hospital status (Hospitalized *vs*. Discharged) of the patient, and the GFI patency (Closed *vs*. Non closed) were recorded in each of the cohort´s examination. Indicators of GFI prognosis thus constructed were correlated demographical, sanitary, surgical and nutritional characteristics of the patients. ***Results***: On conclusion of the study indicators of GIF prognosis behaved as follows: *Mortality*: 14.7 %; *Prolonged hospitalization*: 46.3 %; *Spontaneous closure of GIF*: 36.2 %. Only the type of GIF influenced upon patient´s survival: *ECF*: 87.0 % vs. *EAF*: 82.3 % (Δ = +4.7 %; χ2 = 6.787; p < 0.05). Similarly, in each examination of the cohort, the number of surviving subjects was always greater among those with ECF: After 30 days: ECF: 92.1 % vs. EAF: 83.9 % (Δ = +8.2 %); After 60 days: ECF: 98.1 % vs. EAF: 90.4 % (Δ = +7.7 %; χ2 = 13.764; p < 0.05). ***Conclusions***: Currently, only the type of GIF influences upon survival of the patient.

**Keywords**: Gastrointestinal fistula / Cohort / Mortality / Survival / Hospitalization / Spontaneous closure.

|  | Item No | Recommendation | Page No |
| --- | --- | --- | --- |
| **Title and abstract** | 1 | (*a*) Indicate the study’s design with a commonly used term in the title or the abstract  ***The Authors***: The title of the work reflects the nature of the study described herein. | 1 |
|  |  | (*b*) Provide in the abstract an informative and balanced summary of what was done and what was found  ***The Authors***: A structured Abstract/Summary has been included summarizing key points of the purposes, the design and the results of the study. | 2 |
| Introduction | | | |
| Background/rationale | 2 | Explain the scientific background and rationale for the investigation being reported  ***The Authors***: The rationale, context and background of the research have been presented in the “Introduction” section | 2-3 |
| Objectives | 3 | State specific objectives, including any prespecified hypotheses  ***The Authors***: The supraobjective of the study, and the subsidiary objectives, were presented in the “Introduction” and “Presentation of the Fistula Day Project” sections. Being an observational study, and in the absence of previous data, no hypothesis was put forward. | 3-4 |
| Methods | | | |
| Study design | 4 | Present key elements of study design early in the paper  ***The Authors***: A “Materials and Methods” section is included to describe all the elements of the study design. | 4-5 |
| Setting | 5 | Describe the setting, locations, and relevant dates, including periods of recruitment, exposure, follow-up, and data collection  ***The Authors***: The setting, locations, and relevant dates, including periods of recruitment, exposure, follow-up, and data collection procedures, are described correspondingly in the “Materials and Methods” section. | 4-5 |
| Participants | 6 | (*a*) Give the eligibility criteria, and the sources and methods of selection of participants. Describe methods of follow-up  ***The Authors***: Surveyors supervising the conduction of the study procedures in the participating hospitals were appointed and properly trained. Main researchers maintained close communication with the local surveyors regarding selection and admission of patients, and further follow-up. | 4-5 |
|  |  | (*b*) For matched studies, give matching criteria and number of exposed and unexposed  ***The Authors***: Not relevant (N/R) in lieu of the study design. | N/R |
| Variables | 7 | Clearly define all outcomes, exposures, predictors, potential confounders, and effect modifiers. Give diagnostic criteria, if applicable  ***The Authors***: Outcomes, exposures, predictors, potential confounders, and effect modifiers of the study are described in the “Materials and Methods” sections. | 4-5 |
| Data sources/ measurement | 8* | For each variable of interest, give sources of data and details of methods of assessment (measurement). Describe comparability of assessment methods if there is more than one group  ***The Authors***: Clinical charts, research records and electronic registries were the sources of relevant data as *per* the “Materials and Methods” section of the report. | 4-5 |
| Bias | 9 | Describe any efforts to address potential sources of bias  ***The Authors***: Exclusion criteria were included in the “Materials and Methods” section. Local surveyors were properly trained in the data collection procedures. | 4-5 |
| Study size | 10 | Explain how the study size was arrived at  ***The Authors***: Being an observational study, no size goal was established. | 4-5 |
| Quantitative variables | 11 | Explain how quantitative variables were handled in the analyses. If applicable, describe which groupings were chosen and why  ***The Authors***: Study variables were analysed according with the data type by means of location, dispersion and aggregation statistics. | 4-5 |
| Statistical methods | 12 | (*a*) Describe all statistical methods, including those used to control for confounding  ***The Authors***: Classical statistical methods were used in the data analysis. Subgrouping according with prespecified criteria was done in other to control for confounding factors and in order to meet the study goals. | 4-5 |
|  |  | (*b*) Describe any methods used to examine subgroups and interactions | 4-5 |
|  |  | (*c*) Explain how missing data were addressed  ***The Authors***: Missing data for a particular patient was addressed by means of the “Last Observation Carried Forward” principle, as stated in the “Materials and Methods” section. Percentage of missing data was kept to a minimal by constant communication with local surveyors. | 4-5 |
|  |  | (*d*) If applicable, explain how loss to follow-up was addressed  ***The Authors***: Losses to follow-up were analysed as per the “Intention-to-treat” principle. | 4-5 |
|  |  | (*e*) Describe any sensitivity analyses  ***The Authors***: Not relevant (N/R) in lieu of the study design. | N/R |
| Results | | |  |
| Participants | 13* | (a) Report numbers of individuals at each stage of study- eg numbers potentially eligible, examined for eligibility, confirmed eligible, included in the study, completing follow-up, and analysed  ***The Authors***: Characteristics of the surveyed patients are given in the “Results” section. Being an observation study, and in the absence of previous data, no attempt was made to produce a figure representing the total number of patients assisted for gastrointestinal fistulas in the countries during a particular period of time. | 5-6 |
|  |  | (b) Give reasons for non-participation at each stage  ***The Authors***: No cases were found of refusal to participate. | 5-6 |
|  |  | (c) Consider use of a flow diagram  ***The Authors***: The narrative is clear and fluid. No need for a flow diagram. | 5-6 |
| Descriptive data | 14* | (a) Give characteristics of study participants (eg demographic, clinical, social) and information on exposures and potential confounders  ***The Authors***: Characteristics of the surveyed patients are given in the “Results” section. The study also included descriptive tables for the reader to have a complete picture of the results. | 5-6 |
|  |  | (b) Indicate number of participants with missing data for each variable of interest  ***The Authors***: A table is included in the “Results” section portraying statistical analysis under “Intention-to-treat” and “Analysis –per-protocol” paradigms. | 5-6 |
|  |  | (c) Summarise follow-up time (eg, average and total amount)  ***The Authors***: Follow-up was prespecified as per study design. | 5-6 |
| Outcome data | 15* | Report numbers of outcome events or summary measures over time  ***The Authors***: Outcomes study are presented and described correspondingly in the “Results” section. | 5-6 |
| Main results | 16 | (*a*) Give unadjusted estimates and, if applicable, confounder-adjusted estimates and their precision (eg, 95% confidence interval). Make clear which confounders were adjusted for and why they were included  ***The Authors***: Unadjusted as well as criteria-adjusted outcomes study are presented and described correspondingly in the “Results” section. Complete results are presented for the reader to have a clear picture of the study purposes, objectives, and design. | 5-6 |
|  |  | (*b*) Report category boundaries when continuous variables were categorized  ***The Authors***: When required, continuous variables were stratified/categorized according the study objectives and design. | 5-6 |
|  |  | (*c*) If relevant, consider translating estimates of relative risk into absolute risk for a meaningful time period  ***The Authors***: Given the observational nature of the study, and thus the observed heterogeneity, no attempt was made to produce risk estimates for the study variables. |  |
| Other analyses | 17 | Report other analyses done—eg analyses of subgroups and interactions, and sensitivity analyses  ***The Authors***: Given the observational nature of the study, and thus the observed heterogeneity, no attempt was made to dwell further in other associations and interactions that nonetheless were deemed of no interest for the present time. |  |
| Discussion |  |  |  |
| Key results | 18 | Summarise key results with reference to study objectives  ***The Authors***: Key results of the study are discussed in the corresponding section. | 7-9 |
| Limitations | 19 | Discuss limitations of the study, taking into account sources of potential bias or imprecision. Discuss both direction and magnitude of any potential bias  ***The Authors***: The limitations of the study are also discussed in the study described herein. | 7-9 |
| Interpretation | 20 | Give a cautious overall interpretation of results considering objectives, limitations, multiplicity of analyses, results from similar studies, and other relevant evidence  ***The Authors***: The study results are put into perspective with other global, regional and local data. The study results are in line with previous reports. | 7-9 |
| Generalisability | 21 | Discuss the generalisability (external validity) of the study results  ***The Authors***: The study described herein is the first of four reports attempting to draw a diagnosis of the current management of gastrointestinal fistulas in Latin American countries. Generalisability of the results will be made once all the possible aspects revealed by the project are presented and discussed. A “Future extensions” section is included to make the reader aware of this contingency. | 7-9 |
| Other information |  |  |  |
| Funding | 22 | Give the source of funding and the role of the funders for the present study and, if applicable, for the original study on which the present article is based  ***The Authors***: No external funding was used for completion of this study. |  |
