## Supplementary material for "Outcomes of gastrointestinal fistulas: Results from a multi-continent, multi-national, multi-center cohort": Letter of Approval by the San Javier Hospital Ethics Committee

Guadalajara, Jal. Mex. April 11th 2018

Attention:
**M.D. Humberto Arenas Màrquez. FACS. FASPEN**
**Leader of the Integrated Intestinal Failure Practice Unit**
**Director of the Research Project: Current Status of the Digestive Tube Post-operative Fistula**
Presented.-

Dear M.D. Arenas:

The Ethics Committee of San Javier Hospital has reviewed the research protocol that you send to this committee entitled: Current Status " Of the Post-Operative Fistula of digestive tract; Multicentric, multinational study. DAY OF THE FISTULA ". In said study it is indicated that the day of the protocol will be May 08 of the present year and in which the record will be coordinated for his team under a system designed based on data (REDCAP).

Patients admitted on the day are selected again at 30 and 60 days after their initial registration for the result of their management in that period of time, following the same methodology of the database that are captured by internet.

On the other hand, the hospital that wishes to participate voluntarily is registered and given an access code for the database, with the information obtained, the patient's confidentiality is always maintained.

After the evaluation of the protocol by the Ethics Committee that was approved for its realization in the Hospital San Javier and we wish you the greatest success in the search of the general objective to identify the prevalence of Post-Operative Gastrointestinal Fistula.

Receive a warm greeting and we congratulate you and to your team to develop this type of study through information technology that allows us to measure the problem of this adverse event worldwide and for give us answers for the solution to decrease the risk of This complication and also for the benefit of patients and reduce costs of care for this adverse event in health systems.

Sincerely,

M.D Eduardo Razón Gutiérrez
Director of the Ethics Committee
San Javier Hospital

HOSPITAL SAN JAN JAVIER
Av. Pablo Casals 640
Col. Prados Providencia
44670 Guadalajara, Jal. Mx.
Tel/Fax (33) 3669 0222
www.hospitalsanjavier.com
